# Disrupted ErbB4 splicing with region-specific severity across the cortical visuospatial working memory network in schizophrenia

**DOI:** 10.1101/2025.04.23.25326229

**Authors:** Youjin Chung, H. Holly Bazmi, David A. Lewis, Daniel W. Chung

## Abstract

**Background:** Visuospatial working memory (vsWM) depends on gamma oscillations generated in multiple areas of a cortical network, including dorsolateral prefrontal (DLPFC), posterior parietal (PPC), visual association (V2), and primary visual (V1) cortices. Gamma oscillations require parvalbumin-expressing interneurons (PVI), and deficient gamma power in the vsWM network of schizophrenia (SZ) is thought to arise from lower PVI activity. We previously proposed that PVI activity, and activity-dependent PV levels, are regulated by shifts in splicing of erb-b2 receptor tyrosine kinase 4 (ErbB4) transcripts between major variants that enhance PVI activity and inactive minor variants. Here, we investigated the region-specific pattern of this splicing shift across the vsWM network and its alterations in SZ.

**Methods:** Levels of ErbB4 splice variants and PV mRNA were quantified from 16 pairs of unaffected comparison (UC) and SZ subjects across DLPFC, PPC, V2, and V1.

**Results:** In UC, the major ErbB4 variants showed progressive enrichment relative to minor variants from the rostral to caudal regions. In SZ, this gradient was disrupted by abnormal shifts toward minor variants, with the magnitude of shifts greater in caudal regions. These major-to-minor splicing shifts correlated with lower PV mRNA levels across all regions in SZ.

**Conclusion:** Greater enrichment of major variants in caudal areas suggests spatially regulated molecular mechanism supporting region-specific levels of PVI activity across the vsWM network. In SZ, the region-specific magnitude of splicing shift to minor variants may drive reduced PVI activity throughout the vsWM network, leading to a network-wide dysfunction that contributes to cognitive impairments.

## Introduction

Complex cognitive processes, such as spatial working memory for visual information, or visuospatial working memory (vsWM), depend on oscillating neural activity at gamma frequency (30-100Hz). Earlier studies emphasized the role of gamma oscillations in the dorsolateral prefrontal cortex (DLPFC) for vsWM(1–3), but recent studies have broadened this view, proposing that vsWM depends on gamma oscillations generated in multiple areas of the cortical vsWM network(4, 5). These cortical areas are present along the rostral-to-caudal axis of the brain, and include DLPFC and posterior parietal (PPC), visual association (V2), and primary visual (V1) cortices(4, 5).

Within these cortical areas, gamma oscillations are proposed to arise from a canonical local circuit comprising excitatory pyramidal neurons (PNs) and inhibitory parvalbumin-expressing interneurons (PVIs)(6). In this circuit, PNs provide excitatory drive to PVIs, which in turn provide phasic inhibition to neighboring PNs(7, 8). Following the decay of inhibition from PVIs, PNs fire synchronously to produce neural oscillations at gamma frequency(9–11). Thus, the activity of PVIs is a key element for generating gamma oscillations in each cortical area of the vsWM network.

The activity of PVIs regulates the expression levels of PV mRNA and protein, suggesting that PV levels serve as a molecular marker of PVI activity(12–14). Prior studies have shown that the expression levels of PV mRNA increase across the vsWM network from DLPFC to V1(15–17). Moreover, these regional differences, at least between DLPFC and V1, appear to be due to greater PV mRNA levels per cell rather than differences in the relative proportion of PVIs(18). These findings suggest that PVI activity becomes progressively higher in more caudal regions, creating an ascending gradient of activity along the rostral-to-caudal axis of the cortical vsWM network. Thus, one goal of the present study is to determine if a similar gradient is also present in molecular pathways that regulate PVI activity.

Individuals with SZ exhibit lower gamma power across frontal, parietal, and visual cortices(19–24) and impaired performance during vsWM tasks(25–27). These findings are thought to reflect, at least in part, alterations in PVIs as both PV mRNA and protein levels are lower in the DLPFC in SZ (28–34), and as shown in more recent studies, in the PPC, V2, and V1 as well(15, 35). Thus, a second goal of this study is to determine if the molecular pathways regulating PVI activity are disrupted in the vsWM network of SZ.

A key molecular pathway regulating the activity of PVIs is the neuregulin-1 (NRG-1)-erb-b2 receptor tyrosine kinase 4 (ErbB4) signaling pathway(36, 37). ErbB4 is expressed mainly in PVIs(38, 39) and the activation of ErbB4 by its ligand NRG-1 promotes the formation of excitatory inputs to PVIs(40, 41). Also, the loss of ErbB4 signaling results in fewer excitatory synaptic inputs to, and reduced activity of, PVIs(14, 42). Finally, the ErbB4 signaling pathway has been shown to regulate the generation of cortical gamma oscillations and to affect cognitive function in mouse models(14, 43–45). Thus, the two goals of this study can be addressed by investigating measures of ErbB4 signaling strength across the vsWM network in individuals without or with a diagnosis of SZ.

ErbB4 signaling strength is regulated by alternative splicing of ErbB4 transcripts at the juxtamembrane (JM) and the cytoplasmic (CYT) loci(33). Splicing at the JM locus yields either the JM-a or JM-b variant based on the inclusion of exon 16 or 15b, respectively, whereas the inclusion or exclusion of exon 26 at the CYT locus generates the CYT-1 or CYT-2 variant, respectively(46). Prior studies have established JM-b/CYT-2 as the major splice variant and JM-a/CYT-1 as the minor splice variant based on their relative expression levels(32, 38, 47). These variants differentially regulate the strength of ErbB4 signaling, as the major JM-b/CYT-2 variant exhibits higher tyrosine kinase activity and results in a greater number of excitatory inputs to PVIs compared to the minor JM-a/CYT-1 variant upon NRG-1 stimulation(41). In the DLPFC of SZ, an abnormal shift in major-to-minor ErbB4 splicing has been reported in PVIs(32, 33), and this abnormal shift in ErbB4 splicing was associated with fewer excitatory inputs to PVIs(31). Collectively, these studies suggest that the shift in ErbB4 splicing is an informative surrogate measure of ErbB4 signaling strength, and that SZ is associated with an abnormal shift in ErbB4 splicing that enriches the minor variant in the DLPFC.

Based on these findings, we investigated the shifts in ErbB4 splicing across four cortical areas of the vsWM network to elucidate the molecular basis of the ascending gradient in PVI activity and its alterations in SZ. In 16 pairs of unaffected comparison (UC) and SZ subjects, we first compared the magnitude of shift in ErbB4 splicing between major and minor variants across the DLPFC, PPC, V2, and V1 in UC subjects. Next, we assessed how ErbB4 splicing shift is disrupted across these cortical areas in SZ relative to UC subjects. Finally, we analyzed the relationship between shifts in ErbB4 splicing and PV mRNA levels across all four areas in both subject groups.

## Methods and Materials

### Human Subjects

Brain specimens were collected during standard autopsies at the Allegheny County Office of the Medical Examiner (Pittsburgh, PA) with next-of-kin consent. Independent clinicians provided consensus DSM-IV diagnoses based on structured interviews and review of medical, neuropathology and toxicology records. The same method was used to confirm the absence of psychiatric diagnoses in UC subjects. All procedures were approved by the University of Pittsburgh Committee for Oversight of Research and Clinical Training Involving Decedents and the Institutional Review Board.

The cohort included 16 subject pairs, each with one SZ subject and one UC subject. The detailed demographic, postmortem and clinical characteristics of this cohort are not included in the preprint but are available upon request. These pairs were previously included in studies that examined ErbB4 splice variant levels in the DLPFC of SZ(32) and PV mRNA levels across DLPFC, PPC, V2 and V1 in SZ(15). Accordingly, the current study provides a technical replication of the prior ErbB4 finding in DLPFC of SZ, and of the region-specific PV expression patterns across the vsWM network of SZ. Subjects were perfectly matched for sex and closely matched for age. There were no significant differences between groups in mean age, postmortem interval (PMI), RNA integrity number (RIN), or tissue storage time at −80°C (**Table 1**). Brain pH significantly differed between subject groups (F_1,30_=0.38, *p*=0.024), but the mean difference was 0.2 pH units and of uncertain biological relevance(48).

**Table 1.** Summary characteristics of human subjects used in this study. M male, F female, W white, B black, SD standard deviation, PMI postmortem interval, RIN RNA integrity number.

|  | Unaffected<br>Comparison | Schizophrenia |  |
| --- | --- | --- | --- |
| Sex | 12M/4F | 12M/4F |  |
| Race | 11W/5B | 11W/5B |  |
|  | Mean±SD | Mean±SD | ANOVA |
| Age (years) | 45.1±12.0 | 44.6±10.8 | $F_{1,30}=0.02, p=0.890$ |
| Brain pH | 6.7±0.2 | 6.5±0.3 | $F_{1,30}=0.38, p=0.024$ |
| PMI (hours) | 14.6±6.0 | 13.8±6.6 | $F_{1,30}=0.14, p=0.709$ |
| RIN | 8.3±0.6 | 8.2±0.6 | $F_{1,30}=0.16, p=0.694$ |
| Storage time (months) | 246.6±45.5 | 243.8±44.5 | $F_{1,30}=0.03, p=0.861$ |

### RNA processing

Fresh-frozen tissue blocks from the right hemisphere were sectioned coronally, promptly frozen, and stored at-80°C for each subject. Four cortical regions (DLPFC, PPC, V2, V1) were identified in Nissl-stained sections from the immersion-fixed left hemisphere(49–51), then sampled in the fresh-frozen right hemisphere as described previously(15). In areas where the cortex was sectioned perpendicular to the pial surface, the gray-white matter boundary was carefully marked with a scalpel and photographed. The number of 40μm sections required to obtain approximately 30mm³ of gray matter was then calculated. These sections were cut using a cryostat, and total gray matter was collected in tubes with Trizol reagent (Invitrogen, Carlsbad, CA), ensuring minimal white matter contamination and optimal RNA preservation. Total RNA was then extracted using the RNeasy Lipid Tissue Mini Kit (Qiagen, Valencia, CA), and complementary DNA (cDNA) was synthesized from standardized total RNA dilutions (10 ng/μl) using SuperScript IV VILO Master Mix (Thermo Fisher, Waltham, MA).

### Quantitative polymerase chain reaction (qPCR)

The qPCR was conducted using Power SYBR Green Mastermix (Thermo Fisher) and the QuantStudio 5 Real-Time qPCR system (Applied Biosystems, Foster City, CA). Primer sets for PV, four ErbB4 splice variants (JM-a, JM-b, CYT-1, CYT-2) and pan-ErbB4 used in this study have been previously shown to yield a single band of the predicted size on a gel, >98% amplification efficiency in standard curve analyses, and distinct single products in dissociation curve analyses(32). Reference genes (Beta actin, GAPDH, cyclophilin) were chosen for their stable expression in the DLPFC of SZ compared to UC subjects(32, 33). In this study, we did not find any statistically significant effect of diagnosis (*F*_1,114_=2.18, *p*=0.143), region (*F*_3,114_=2.48, *p*=0.064), or diagnosis-by-region interaction (*F*_3,114_=0.10, *p*=0.961) on the geometric mean of beta actin, GAPDH, and cyclophilin expression levels.

To minimize experimental variability, cDNA samples from each region of the matched subject pairs were amplified on the same 386-well plate. The difference in CT values (dCT) for each transcript was calculated by subtracting the geometric mean CT of the reference genes from the target transcript’s CT value. Transcript levels were determined as expression ratios (2^-dCT^). Consistent with our previous studies(32, 33), ErbB4 splice variant levels were normalized to pan-ErbB4 levels to reduce variability due to differences in total ErbB4 expression across subjects.

### Statistics

A repeated measures model was implemented in SAS PROC MIXED using the REML method to analyze region-specific expression of ErbB4 splice variants and PV transcripts as previously described(15, 32). In the REML method, the containment method was used to compute denominator degrees of freedom. To assess the effect of cortical regions on transcript levels in the UC group, the model included transcript level as a dependent variable, cortical region as a fixed effect, and sex, age, PMI, storage time, RIN, and brain pH as covariates. To assess the effect of diagnosis on transcript levels in each cortical area, the model included diagnosis, region, and their interaction as fixed effects, with sex, age, PMI, storage time, RIN, and brain pH as covariates. Main effects were tested using F-tests, and post hoc tests used the Holm method to correct for multiple comparisons. The 16 subject pairs in this study are a subset of a larger cohort previously used to examine ErbB4 splice variant expression levels in the DLPFC(32). Accordingly, for the analyses assessing the region effect in UC group, the DLPFC served as a reference region for post hoc comparisons as findings in this region represented a technical replication of the prior work.

A composite ErbB4 splicing score was obtained by first computing the ratio of major to minor variant levels for each splice locus. Next, Z-scores were calculated for the log-transformed variant ratio at each locus, considering all regions and all subjects. The composite splicing score for each region for a given subject was calculated by averaging the Z-scores for JM and CYT loci and then normalizing to the minimum splicing score across all regions and subjects.

The impact of co-occurring factors common in SZ, such as schizoaffective diagnosis, the presence of a substance use disorder at the time of death, use of nicotine, antidepressants, benzodiazepines, or valproic acid at the time of death, or death by suicide was evaluated using ANCOVA models. Each factor and region were included as main effects, with age, sex, PMI, brain pH, RIN, and storage time as covariates.

To examine relationships between variables across cortical regions while accounting for within-subject dependencies, the repeated measures correlation was performed using the rmcorr package in R as previously described(52). This approach estimates a shared within-subject regression slope (R_rm_) for repeated measures across multiple subjects, effectively capturing intra-individual associations while controlling for inter-individual variability.

## Results

### Expression levels of Pan-ErbB4 in the vsWM network of unaffected and schizophrenia subjects

We first examined the overall expression of Pan-ErbB4 across the DLPFC, PPC, V2, and V1 in 16 pairs of UC and SZ subjects. The mixed model did not detect any significant effect of region, diagnosis, or diagnosis-by-region interaction (**Figure S1**). Thus, for the subsequent analyses of ErbB4 splice variants, we normalized their levels to Pan-ErbB4 to reduce variability arising from non-significant differences in the total ErbB4 expression between subjects.

### ErbB4 splicing shift between major and minor variants in the vsWM network of unaffected subjects

Next, we examined the normalized levels of major and minor variants of ErbB4 transcripts across the DLPFC, PPC, V2, and V1 from UC subjects. Although the mixed model did not detect a significant region effect on normalized JM-a levels, these levels were lower in each of the other areas compared to DLPFC with the effect size smallest in PPC, intermediate in V2, and largest in V1 (**Figure 1A**). The mixed model also did not detect a significant region effect on normalized JM-b levels, with small effect sizes for higher JM-b levels in each of the other three regions relative to DLPFC (**Figure 1B**). Next, the mixed model showed a significant region effect on the normalized CYT-1 levels, with the post hoc analyses revealing significantly lower CYT-1 levels in V2 and V1 compared to DLPFC (**Figure 1C**). Furthermore, the effect sizes for lower CYT-1 levels relative to DLPFC progressively increased from PPC, to V2, and then to V1. Finally, the mixed model detected a significant region effect on normalized CYT-2 levels (**Figure 1D**), but the post hoc analyses revealed no significant differences in CYT-2 levels in PPC, V2, or V1 relative to DLPFC.

**Figure 1.**
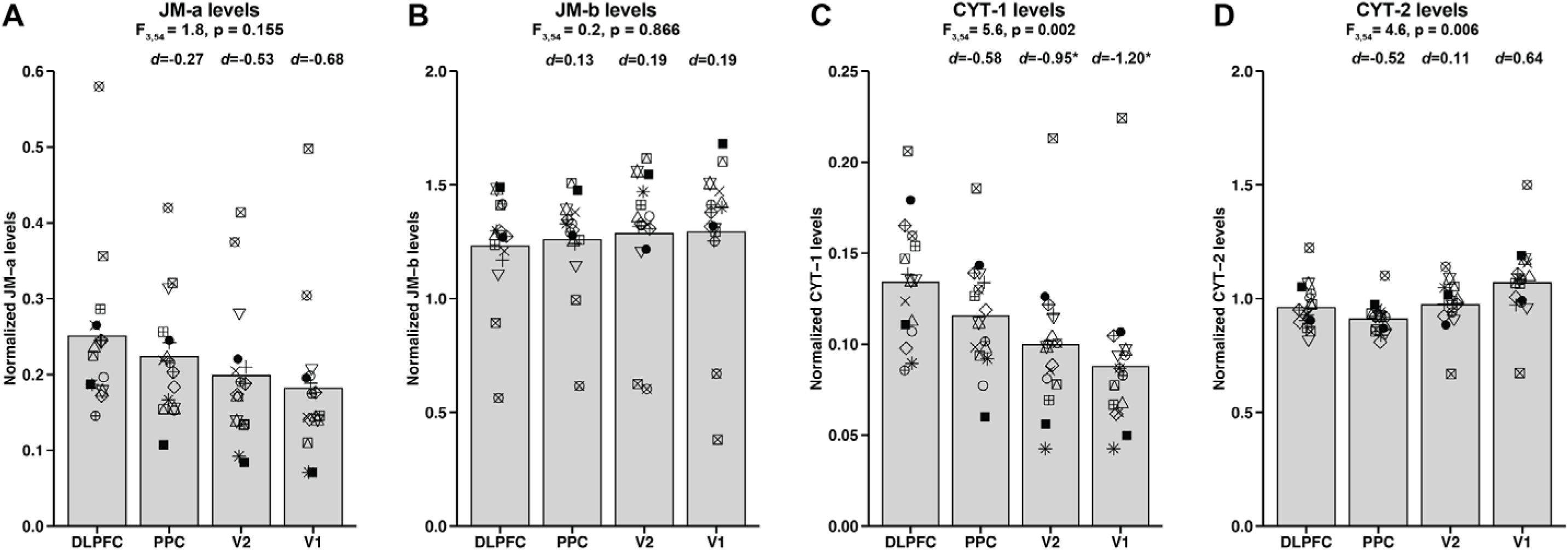
ErbB4 splice variant expression in the vsWM network of unaffected comparison subjects. Bar graphs comparing expression levels of the ErbB4 JM-a (**A**), JM-b (**B**), CYT-1 (**C**), and CYT-2 (**D**) splice variants across DLPFC, PPC, V2, and V1 of UC subjects. Bars represent the group mean and each subject is represented by the same unique symbol across the four regions. Numbers above each bar represent the effect size of each dependent variable relative to DLPFC. The effect size for lower JM-a levels relative to DLPFC was smallest in PPC, intermediate in V2, and greatest in V1 (**A**), while JM-b levels showed small effect sizes for higher expression in PPC, V2, and V1 relative to DLPFC (**B**). The effect size for lower CYT-1 levels relative to DLPFC progressively increased from PPC to V2 and V1 (**C**), whereas CYT-2 showed a significant region effect but no consistent effect size pattern relative to DLPFC (**D**). Asterisks indicate significant difference compared to DLPFC after multiple comparison correction (*p*<0.05).

To compare the relative shift between major and minor variant levels across the DLPFC, PPC, V2, and V1 from UC subjects, we assessed the ratio of JM-b to JM-a levels (i.e., JM-b:JM-a ratio) and CYT-2 to CYT-1 levels (i.e., CYT-2:CYT-1 ratio). The mixed model revealed a significant region effect on the JM-b:JM-a ratio, with post hoc analyses showing a higher ratio in V1 compared to DLPFC. The effect size for higher JM-a:JM-b ratio relative to DLPFC was smallest in PPC, intermediate in V2, and largest in V1 (**Figure 2A**). Similarly, the mixed model showed a significant region effect on the CYT-2:CYT-1 ratio, with post hoc analyses indicating significantly higher ratios in V2 and V1 compared to DLPFC (**Figure 2B**). The effect size for higher CYT-2:CYT-1 ratio relative to DLPFC was smallest in PPC, intermediate in V2, and largest in V1. Finally, there was a strong positive repeated measures correlation between the JM[1]b:JM[1]a and CYT[1]2:CYT[1]1 ratios across cortical regions (**Figure 2C**). Together, these findings indicate greater enrichments of major ErbB4 variants in the caudal compared to rostral regions of the vsWM network and that these splicing shifts occur in tandem at the JM and CYT loci.

**Figure 2.**
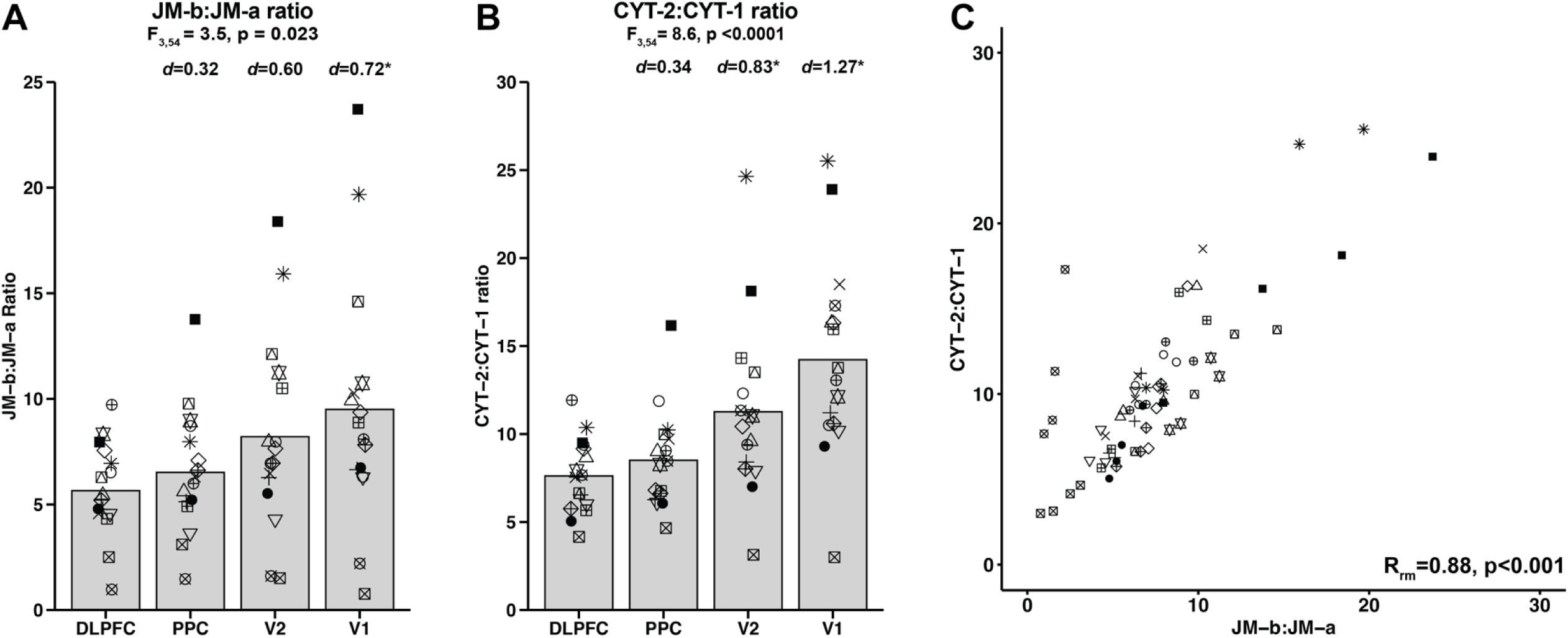
Splice variant ratio in the vsWM network of unaffected comparison subjects. (**A, B**) Bar graphs comparing the JM-b:JM-a ratio (**A**) and the CYT-2:CYT-1 ratio (**B**) across DLPFC, PPC, V2, and V1 of UC subjects. Bars represent the group mean and each subject is represented by the same unique symbol across the four regions. Numbers above each bar represent the effect size of each dependent variable relative to DLPFC. Asterisks indicate significant difference compared to DLPFC after multiple comparison correction (*p*<0.05). From DLPFC to V1, the splice variant ratios show a progressively increasing trend, indicating greater enrichment of major variants relative to minor variants in the caudal compared to rostral regions. (**C**) Repeated measures correlation (R_rm_) plot between the JM-b:JM-a ratio (x-axis) and the CYT-2:CYT-1 ratio (y-axis) across DLPFC, PPC, V2 and V1. Each subject is represented by the same unique symbol across all regions. R_rm_ value indicates the estimated within-subject regression slope for repeated measures across all UC subjects. A strong positive R_rm_ value suggests that the shifts in splicing at the JM and CYT loci occur in tandem across the four regions of the vsWM network.

### ErbB4 splicing shift between major and minor variants in the vsWM network of schizophrenia subjects

We next investigated whether the JM-b:JM-a and CYT-2:CYT-1 ratios across regions of the vsWM network are altered in SZ. For JM-b:JM-a ratio, the mixed model showed significant effects of diagnosis and diagnosis-by-region interaction (**Figure 3A**). Post hoc analyses further indicated significantly lower JM-b:JM-a ratios in each of the four cortical regions in SZ. Additionally, the effect sizes for lower JM-b:JM-a ratio in SZ progressively increased along the rostral-to-caudal axis. For CYT-2:CYT-1 ratios, the mixed model revealed significant effects of diagnosis, region, and diagnosis-by-region interaction (**Figure 3B**). Post hoc analyses revealed significantly lower CYT-2:CYT-1 ratios in the four cortical regions in SZ, with the effect size progressively increasing along the rostral-to-caudal axis. Finally, there was a strong positive repeated measures correlation between the within-pair differences in the CYT-2:CYT-1 ratio and the corresponding differences in the JM-b:JM-a ratio across cortical regions (**Figure 3C**). Together, these findings indicate greater enrichments of minor variants in the caudal compared to rostral regions in SZ relative to UC and that these splicing shifts occur in tandem at the JM and CYT loci.

**Figure 3.**
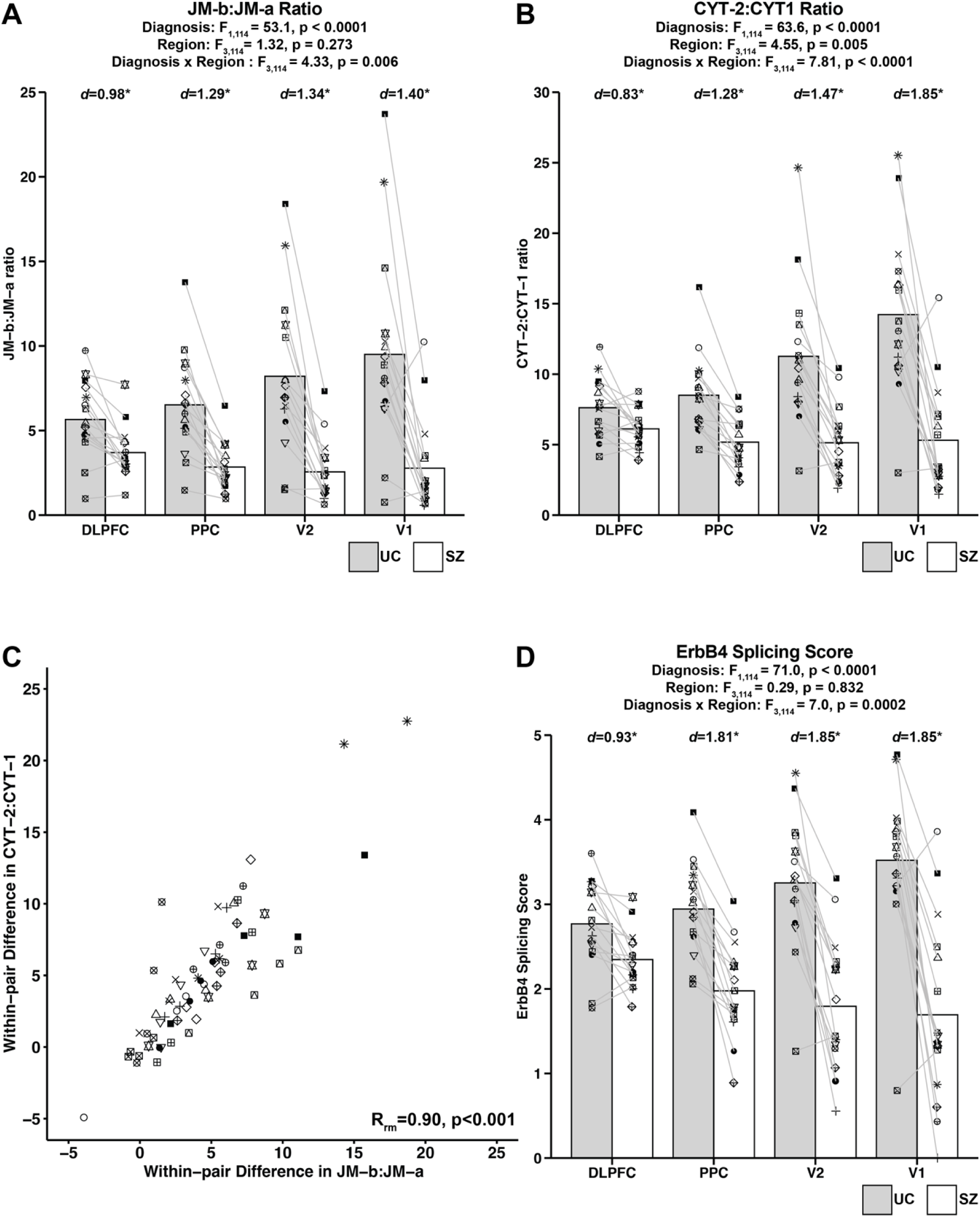
Abnormal ErbB4 splicing shift in the vsWM network of schizophrenia subjects. (**A, B, D**) Bar graphs comparing the JM-b:JM-a ratio (**A**), CYT-2:CYT-1 ratio (**B**), and the composite ErbB4 splicing score (**D**) between UC (gray bar) and SZ (white bar) subjects across DLPFC, PPC, V2, and V1. Bars represent the group mean and each subject is represented by the same unique symbol across the four regions. Numbers above each bar represent the effect size within each region in SZ relative to UC subjects. Asterisks indicate significant differences between UC and SZ subjects after multiple comparison correction (*p*<0.05). Splice variant ratios (**A, B**) and the splicing scores (**D**) are lower across all four regions in SZ with the magnitude of deficit progressively increasing from DLPFC to V1, indicating greater abnormal enrichment of minor variants in the caudal compared to rostral regions. (**C**) Repeated measures correlation (R_rm_) plot between the within-pair differences in the JM-b:JM-a ratio (x-axis) and in the CYT-2:CYT-1 ratio (y-axis) across DLPFC, PPC, V2 and V1. Each subject pair is represented by the same unique symbol across the regions. R_rm_ value indicates the estimated within-subject pair regression slope for repeated measures across all pairs. A strong positive R_rm_ value suggests that the abnormal splicing shifts at the JM and CYT loci occur in tandem across the four regions of the vsWM network in SZ relative to UC subjects.

### Composite analysis of ErbB4 splicing shifts in the vsWM network of schizophrenia subjects

Given that splicing changes in the JM and CYT loci likely occur in tandem, we calculated the composite score of ErbB4 splicing shift by averaging Z-scores of log-transformed variant ratios at each locus (see Methods). This composite splicing score, where a lower score indicates a greater enrichment of the minor variant, was used to investigate the overall shift in ErbB4 splicing in the vsWM network of SZ. The mixed model showed significant effects of diagnosis and diagnosis-by-region interaction on the splicing score, and the post hoc analyses showed significantly lower composite splicing scores in SZ across the DLPFC, PPC, V2 and V1 (**Figure 3D**). Furthermore, the effect size for lower splicing score in SZ was smallest in DLPFC, intermediate in PPC, and largest in both V2 and V1. Finally, The ErbB4 splicing score did not significantly differ as a function of a diagnosis of schizoaffective disorder; the presence of a substance use disorder at the time of death; use of nicotine, antidepressants, benzodiazepines, or valproic acid at the time of death; or death by suicide (**Figure S2**), suggesting this alteration reflects the disease process of SZ, and not the presence of common co-occurring factors.

### Relationship between ErbB4 splicing shift and PV mRNA levels in the vsWM network of unaffected and schizophrenia subjects

To investigate the relationship between ErbB4 splicing shift and PV mRNA levels in the vsWM network, we first measured PV mRNA levels across the DLPFC, PPC, V2, and V1 from UC subjects. The mixed model revealed a significant region effect on PV mRNA levels, with the post hoc analyses showing higher levels in PPC, V2 and V1 compared to DLPFC. Also, the effect size for higher PV mRNA levels relative to DLPFC was smallest in PPC, intermediate in V2, and largest in V1 (**Figure 4A**). Moreover, there was a strong positive repeated measures correlation between the ErbB4 splicing score and PV mRNA levels across cortical regions (**Figure 4B**), suggesting that greater enrichment of the major ErbB4 variant is associated with higher PV mRNA levels in the vsWM network.

**Figure 4.**
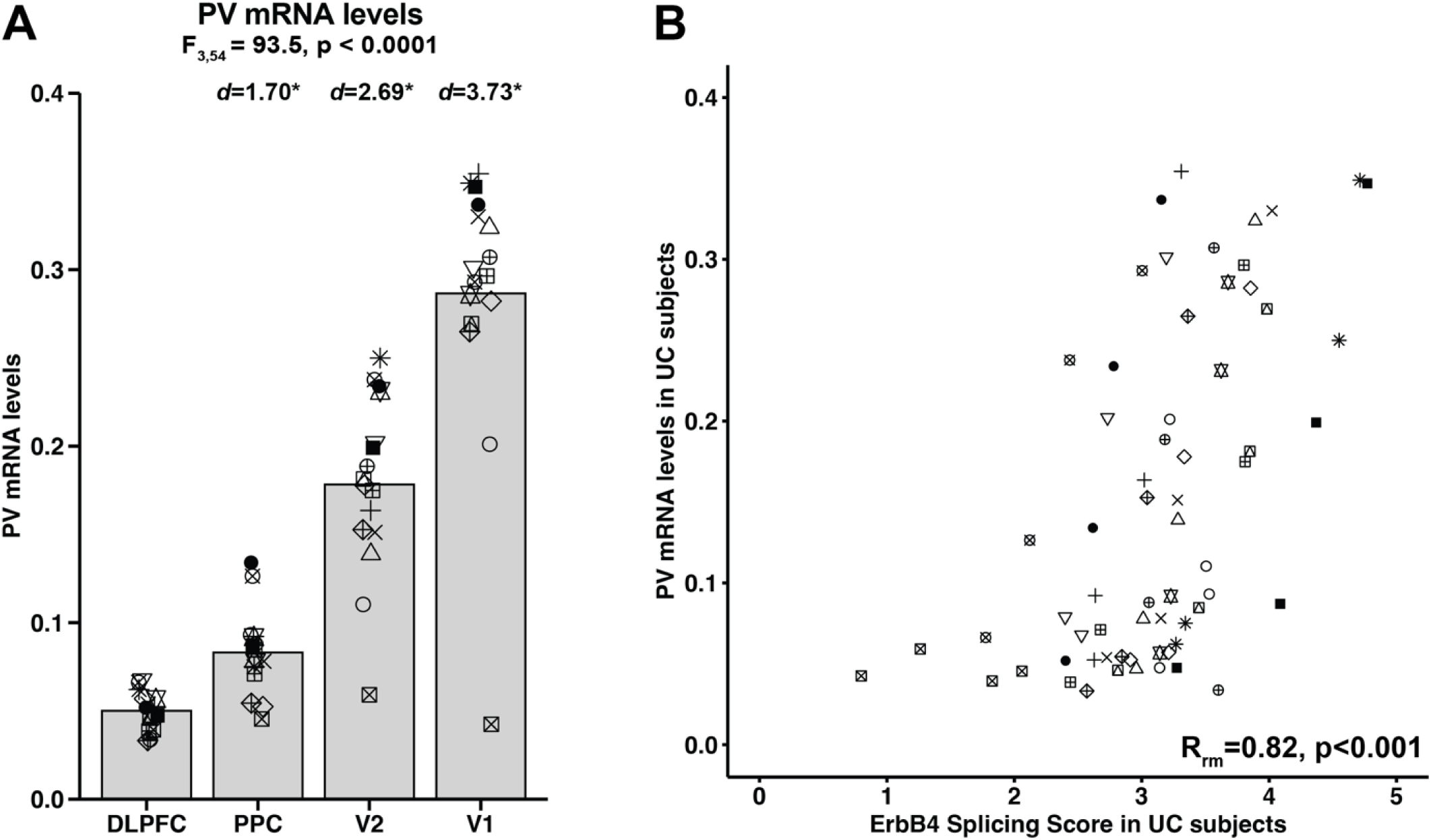
Relationship between shift in ErbB4 splicing and PV mRNA levels in the vsWM network of unaffected comparison subjects. (**A**) Bar graph comparing PV mRNA levels across DLPFC, PPC, V2, and V1 of UC subjects. Bars represent the group mean and each subject is represented by the same unique symbol across the four regions. Numbers above each bar represent the effect size relative to DLPFC. Asterisks indicate significant differences compared to DLPFC after multiple comparison correction (*p*<0.05). Similar to ErbB4 splicing score, PV mRNA level shows a progressively increasing trend from DLPFC to V1. (**B**) Repeated measures correlation (R_rm_) plot between the composite ErbB4 splicing score (x-axis) and PV mRNA levels (y-axis) across DLPFC, PPC, V2 and V1 of UC subjects. Each subject is represented by the same unique symbol across all regions. R_rm_ value indicates the estimated within-subject regression slope for repeated measures across all UC subjects. A strong positive R_rm_ value suggests that greater enrichment of the major ErbB4 variant is associated with higher PV mRNA levels in the vsWM network.

Next, we investigated whether PV mRNA levels in regions of the vsWM network were altered in SZ. The mixed model showed significant effects of diagnosis, region, and diagnosis-by-region interaction (**Figure 5A**). The post hoc analyses further showed significantly lower PV mRNA levels in SZ in every region, with the effect size increasing along the rostral-to-caudal axis. Furthermore, there was a strong positive repeated measures correlation between the within-pair differences in ErbB4 splicing scores and the within-pair differences in PV mRNA levels across cortical regions (**Figure 5B**). To further validate this relationship, we assessed whether the alterations in ErbB4 splicing scores and PV mRNA levels in SZ occurred in the same direction. Among the 64 observations derived from 16 subject pairs across four brain regions, 56 observations exhibited concordant changes in ErbB4 splicing score and PV mRNA levels, with 52 showing lower values (upper right quadrant in **Figure 5B**) and 4 showing higher values (lower left quadrant in **Figure 5B**) for both measures in SZ. Finally, a binomial test confirmed that the number of concordant observations (56 out of 64) is significantly greater than expected by chance (p < 0.0001). Together, these findings suggest that greater enrichment of minor ErbB4 variant is associated with larger deficits in PV mRNA levels across the vsWM network in SZ.

**Figure 5.**
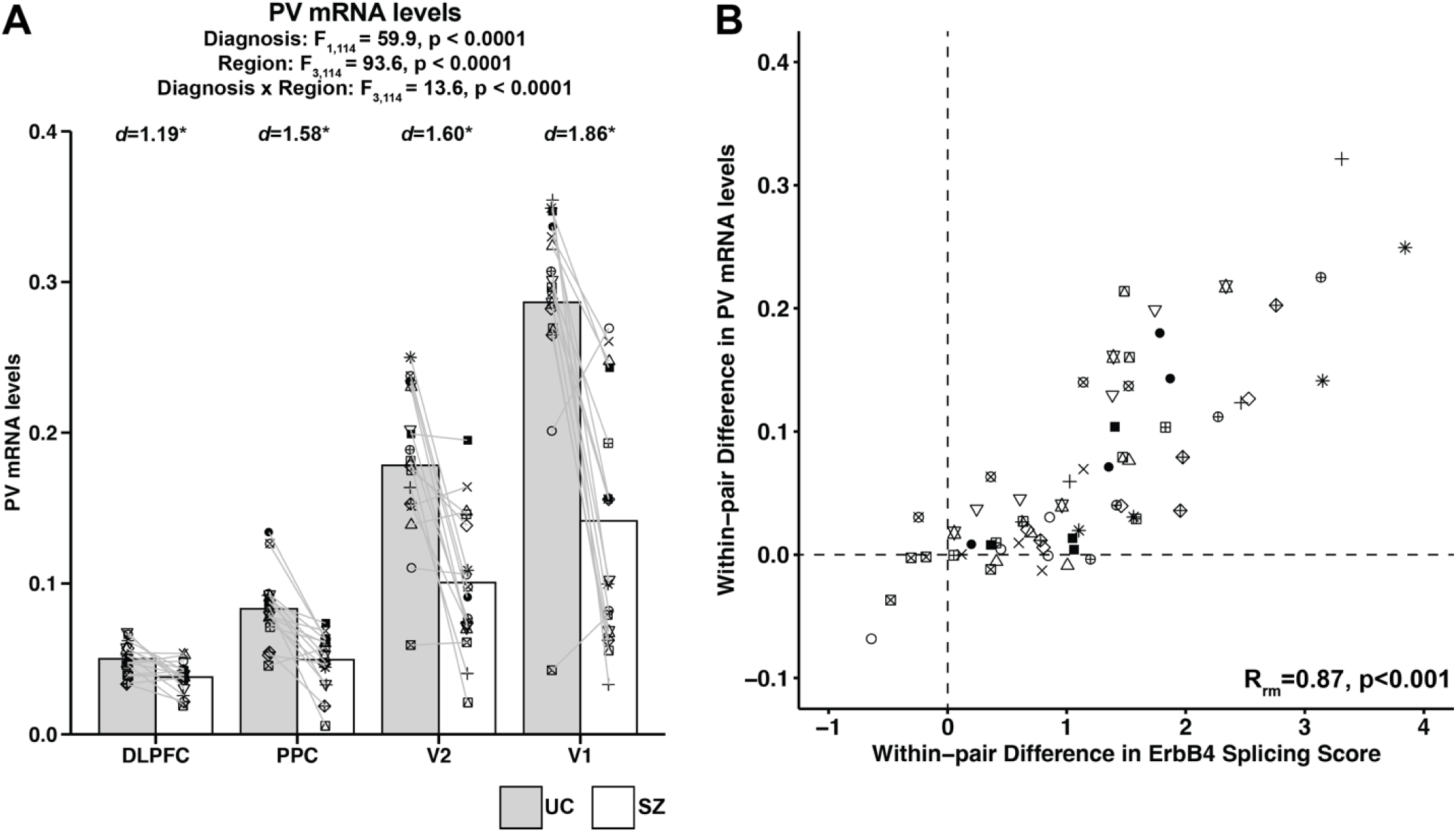
Relationship between abnormal shift in ErbB4 splicing and deficits in PV mRNA levels in the vsWM network of schizophrenia subjects. (**A**) Bar graph comparing PV mRNA levels between UC (gray bar) and SZ (white bar) subjects across DLPFC, PPC, V2, and V1. Bars represent the group mean and each subject is represented by the same unique symbol across the four regions. Numbers above each bar represent the effect size of each dependent variable in SZ relative to UC subjects. Asterisks indicate significant differences in SZ compared to UC subjects after multiple comparison correction (*p*<0.05). PV mRNA levels are lower across all four regions in SZ with the magnitude of their deficits progressively increasing from DLPFC to V1. (**B**) Repeated measures correlation (R_rm_) between the within-pair differences in the ErbB4 splicing score (x-axis) and in the PV mRNA levels (y-axis) across DLPFC, PPC, V2 and V1. Each subject pair is represented by the same unique symbol across the regions. R_rm_ value indicates the estimated within-subject pair regression slope for repeated measures across all pairs. A strong positive R_rm_ value suggests that greater abnormal enrichment of minor ErbB4 variant is associated with larger magnitude of deficits in PV mRNA levels across the vsWM network in SZ. The upper right and lower left quadrants, divided by black dotted lines, represent observations where both ErbB4 splicing scores and PV mRNA levels were altered in the same direction (56 out of 64 observations.).

## Discussion

In this study, we determined the region-specific pattern of ErbB4 splice variant levels across the four cortical regions of the vsWM network—DLPFC, PPC, V2, and V1—in UC subjects, and investigated whether ErbB4 splicing is shifted across these regions in SZ. In UC subjects, the ratio of major to minor ErbB4 splice variants was progressively higher along the rostral-to-caudal axis, with the highest ratio observed in V1. In SZ subjects, this ratio was lower across all regions, with larger decrements in the caudal areas. These findings suggest that ErbB4 splicing follows a rostral-to-caudal gradient in UC subjects, with greater enrichment of major variants in caudal regions. In SZ, this gradient is disrupted by a shift toward minor variants, with the alteration more pronounced in caudal regions.

Alternative splicing of ErbB4 transcripts determines the level of ErbB4 kinase activity, with the major variants exhibiting higher tyrosine kinase activity compared to the minor variants(41). This kinase activity promotes the formation of excitatory synaptic inputs to PVIs(14, 40, 41), suggesting that the balance of ErbB4 splice variants may serve as a key determinant of PVI activity and thus of activity-dependent PV levels. In UC subjects, PV mRNA levels progressively increased along the rostral-to-caudal axis, suggesting higher PVI activity in caudal regions of the vsWM network. Moreover, these higher PV levels were associated with greater enrichment of major ErbB4 variants across cortical regions, supporting the idea that ErbB4 splicing modulates regional differences in PVI activity. In SZ subjects, PV mRNA levels were significantly lower across all regions, with the magnitude of reduction progressively increasing from rostral to caudal areas of the vsWM network. Furthermore, greater decreases in PV mRNA levels were associated with greater shifts toward the minor ErbB4 splice variants, suggesting that disruption of ErbB4 splicing may contribute to regionally graded reductions in PVI activity in the illness. Together, these findings suggest that spatially regulated ErbB4 splicing serves as a conserved mechanism for modulating PVI activity across the vsWM network, and that its disruption in SZ may contribute to the network-wide deficits in PVI activity.

The strength of our findings is reinforced by their consistency with prior results. First, the current study replicated the previously reported major-to-minor shift in ErbB4 splicing in the DLPFC of SZ(32, 33). Second, the current study replicated the prior finding of progressively increasing PV mRNA levels from DLPFC to V1 in UC subjects and a greater magnitude of PV deficits along the same axis in SZ(15). Furthermore, the current study replicated our previous finding that the major-to-minor shift in ErbB4 splicing predicts PV mRNA levels in the DLPFC of SZ(32, 33). Finally, the present study broadens the spatial scope of this relationship by demonstrating its presence across multiple cortical regions in both UC and SZ subjects, suggesting that ErbB4 splicing may play a network-wide role in regulating PVI activity throughout the vsWM system.

Several limitations are important to consider in interpreting the results of this study. First, while ErbB4 is predominantly expressed in PVIs, it is also present in calretinin-expressing interneurons (CRIs)(38, 39). Because our qPCR analyses were conducted on total gray matter, the measured changes in ErbB4 splicing may not be exclusively from PVIs but could also be from CRIs. However, our prior study found no significant alterations in ErbB4 splicing in layer 2 of the DLPFC of SZ where CRIs are predominantly localized(32), suggesting that ErbB4 splicing is largely unaffected in CRIs in the DLPFC. Furthermore, the DLPFC is the most rostral area examined in this study, and CRIs are more abundant in rostral than caudal regions(53). Thus, the ErbB4 splicing deficits observed in cortical regions caudal to DLPFC in this study are also unlikely to originate from CRIs. Second, analyses of individual splice variant levels showed limited and inconsistent region effects across the vsWM network in UC subjects.

For example, the mixed model did not detect a significant region effect for JM-a or JM-b levels, and although CYT-2 levels showed a significant region effect, they did not exhibit any rostral-to-caudal gradient pattern. However, the splice variant ratios showed both a significant region effect and a consistent rostral-to-caudal gradient, likely because these ratios capture the reciprocal changes in major and minor variants that occur during splicing shift. Thus, the splice variant ratios represent a more sensitive measure of ErbB4 splicing shifts across the vsWM network than individual variant levels alone.

We observed concordant changes in ErbB4 splicing across the JM and CYT loci in both UC and SZ subjects, with a strong positive relationship between the shifts in splicing ratios at these loci. These coordinated changes suggest that common upstream regulatory mechanisms may influence splicing at both loci in the vsWM network. Furthermore, while the coordinated splicing pattern is preserved in SZ, the abnormal shifts toward minor variants at both loci indicate a disruption in these common regulatory mechanisms. Prior studies suggest that these regulatory mechanisms may involve RNA-binding proteins such as ELAVL2 and PTBP2, which regulate ErbB4 splicing at the JM and CYT loci, respectively(54, 55). Thus, future studies should investigate if changes in the levels and/or function of these RNA-binding proteins contribute to the cortical gradient of ErbB4 splicing and its region-specific disruption in the vsWM network of SZ.

In conclusion, our study suggests that ErbB4 splicing could serve as a molecular mechanism for determining different levels of PVI activity across cortical regions of the vsWM network. Furthermore, the disruption of this mechanism may contribute to the region-specific reductions in PVI activity, thereby impairing the generation of gamma oscillations across the vsWM network in SZ. Thus, our study provides key insight into how molecular alterations with varying severity across distinct cortical areas may drive network-wide dysfunction underlying cognitive impairments in the illness.

## Data Availability

All data produced in the present study are available upon reasonable request to the authors

## Acknowledgments

This work was supported by the Rising Star Fund from the University of Pittsburgh Department of Psychiatry and National Institute of Health grant 5T32MH016804 (to D.W.C.), and by National Institutes of Health grants MH103204 and MH043784 (to D.A.L.).

