## Supplemental Data for "Disrupted ErbB4 splicing with region-specific severity across the cortical visuospatial working memory network in schizophrenia"

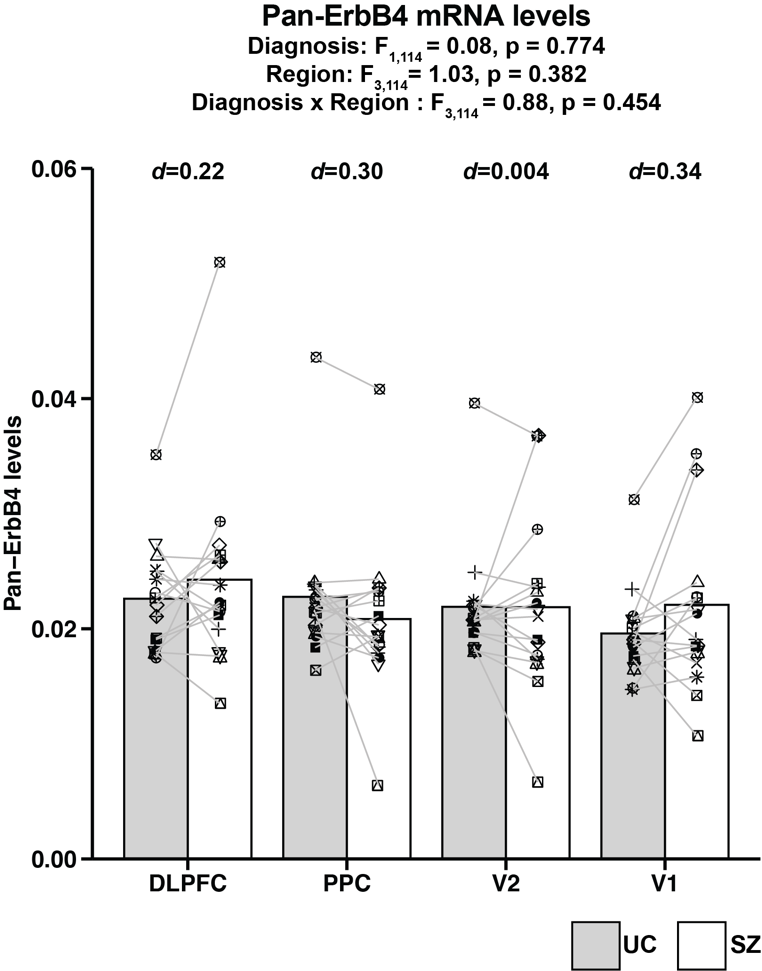

**Figure S1. Pan-ErbB4 mRNA levels in the vsWM network of unaffected comparison and schizophrenia subjects.** Bar graph comparing the Pan-ErbB4 mRNA levels between UC (gray bar) and SZ (white bar) subjects across DLPFC, PPC, V2, and V1. Bars represent the group mean and each subject is represented by the same unique symbol across the four regions. Numbers above each bar represent the effect size within each region in SZ relative to UC subjects.

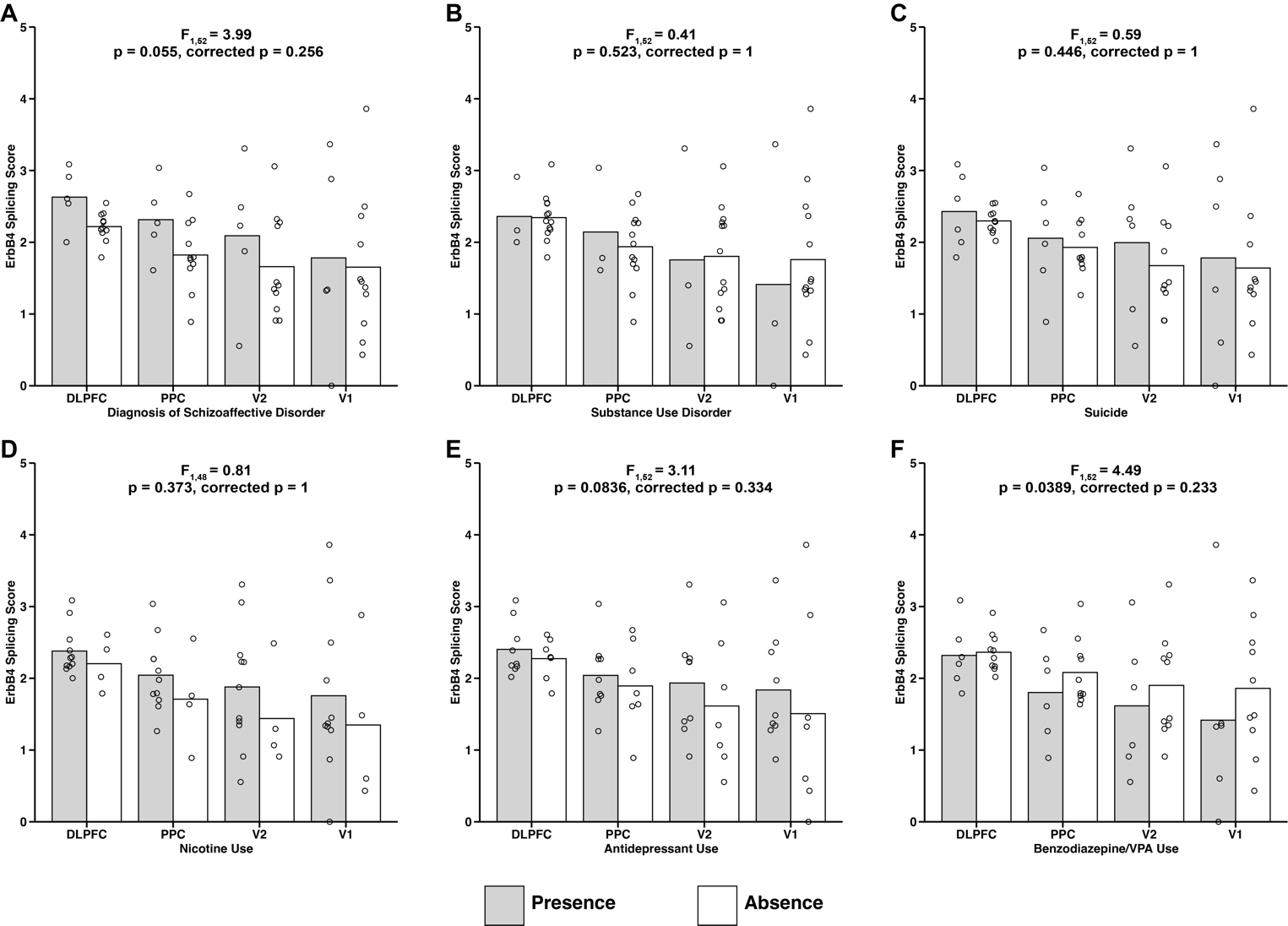

**Figure S2. Lack of effect from common co-occuring factors on the composite ErbB4 splicing score in schizophrenia subjects.** Bar graphs showing the ErbB4 splicing scores in SZ subjects across DLPFC, PPC, V2 and V1 grouped by presence (gray bar) or absence (white bar) of each co-occuring factor commonly associated with the illness. Circles represent individual subject. Statistics from ANCOVA model and the Holm-corrected p-values for the effect of each factor are shown above each graph.
